# Association of *SAPAP3* allelic variants with symptom dimensions and pharmacological treatment response in obsessive-compulsive disorder

**DOI:** 10.1101/2020.03.07.20032391

**Authors:** Safoora Naaz, Srinivas Balachander, Nithyananda Srinivasa Murthy, MS Bhagyalakshmi, Reeteka Sud, Priyanka Saha, Janardhanan C Narayanaswamy, YC Janardhan Reddy, Sanjeev Jain, Meera Purushottam, Biju Viswanath

## Abstract

**Background:** Though several *SAPAP3* gene knockout studies in mice have implicated its role in compulsivity, human studies have failed to demonstrate its association with obsessive-compulsive disorder (OCD). We examined the association between allelic variants of a single nucleotide polymorphism in the *SAPAP3* gene (rs6662980) with specific aspects of the OCD phenotype.

**Methods:** A total of 200 subjects with OCD were genotyped using the TaqMan assay. All subjects were assessed using Mini International Neuropsychiatric Interview, the Yale-Brown Obsessive-Compulsive Scale, and their treatment response was evaluated over naturalistic treatment and follow-up.

**Results:** After correcting for multiple comparisons, G-allele at rs6662980 was found to be associated with contamination/washing symptoms (p=0.003). Logistic regression analysis also showed that presence of G allele predicted poor response to serotonin reuptake inhibitors [odds ratio = 2.473 (95% CI = 1.157 - 5.407), p=0.021]. Interaction between presence of G-allele and contamination factor score predicted SRI resistance (B= 1.197, p = 0.006).

**Limitations:** We did not use a dimensional measure for assessing OCD symptoms. Treatment response was assessed over naturalistic follow-up.

**Conclusion:** Specific phenotypic manifestations of OCD, which include contamination and washing-related symptoms along with resistance to serotonin reuptake inhibitors, may be related to alterations in the *SAPAP3* gene.

**Public Significance Statement:** “This study finds that a specific polymorphism in the SAPAP3 gene, was found to be associated with the contamination/washing symptoms of OCD and was also found to predict resistance to pharmacological treatment”

**Highlights:**

- *SAPAP3* gene is implicated in OCD
- Association of *SAPAP3* gene (rs6662980) with OCD phenotypes was examined
- Presence of the minor (G) allele predicted contamination & washing symptoms
- *SAPAP3* polymorphism had a significant association with treatment response
- Interaction between presence of G-allele and contamination factor score predicted treatment resistance

## 1. Introduction

Obsessive-compulsive disorder (OCD) is a severe, chronic and debilitating neuropsychiatric disorder, that affects ∼2 % of the world population (Ruscio et al., 2010). OCD has a strong genetic basis, with heritability estimates ranging from 27-47% (adult-onset) to 45-65% (childhood-onset) (Van Grootheest et al., 2007). Several candidate-gene and genome-wide association studies in OCD have implicated the role of genes in serotonergic, dopaminergic and glutamatergic systems. However, very few of these findings have been consistently replicated. (Pauls et al., 2014).

Neuroimaging, genetic and neurochemical evidence, as well as insights from animal models, suggest that glutamatergic signaling is dysregulated in OCD (Pittenger et al., 2011). Many genes in the glutamatergic system like *GRIN2B, SAPAP3* and *SLC1A1* have been associated with OCD (Pauls, 2010). Pharmacologic agents modulating glutamatergic transmission such as memantine, topiramate and riluzole have shown some promise in the treatment of OCD (Grados et al., 2013). Further, glutamatergic function is also altered by serotonin reuptake inhibitors (SRIs), which are the first line treatment for OCD (Ohno, 2018; Rodrigues et al., 2015).

Mice with mutated versions of *SAPAP3*, (also *DLGAP3*) exhibit OCD-like behavior, including excessive grooming, increased anxiety and abnormal cortico-striatal neurotransmission which was significantly reduced after Fluoxetine treatment (Burguière et al., 2013; Manning et al., 2019; Welch et al., 2007). SAPAP3 belongs to highly homologous SAPAP family proteins, which are components of the post-synaptic density that interact with multiple members of proteins from PSD95- and SHANK-families, to form a postsynaptic scaffolding complex, which plays an important role in facilitating organization of post synaptic signaling at the glutamatergic synapse (Scannevin and Huganir, 2000).

SAPAP3, the only member which is highly expressed in the striatum, has been studied in OCD spectrum disorders. A family-based association study showed *SAPAP3* as a promising functional candidate gene for human grooming disorder (Boardman et al., 2011). Another case-control study supports the role of *SAPAP3* in Trichotillomania (TTM) and OCD (Züchner et al., 2009). As OCD is a heterogeneous disorder, it may be important to study the association of its specific phenotypic manifestations with genetic factors. A study in the South African population links *SAPAP3* variants to the development of early onset OCD (Boardman et al., 2011).

Despite several replications in mice models which have shown *SAPAP3* to be associated with compulsive behaviors, none of the human genome-wide studies done in OCD have been able to detect significant SNPs in this region. The previously mentioned family-based candidate gene study (Bienvenu et al., 2009) which examined multiple SNPs within the *SAPAP3* gene found the presence of G allele in rs6662980 to be nominally associated with Grooming Disorders (relative risk = 1.6). In the db-SNP database, the minor allele frequency of this variant in South Asian populations is reported to be 0.21 (1000Genomes) and 0.23 (PAGE) (Sherry et al., 2001). In this current study, we examine the association between clinical characteristics of OCD (symptom dimensions, disease severity, age at onset, comorbidity, treatment response) and an SNP in the *SAPAP3* gene (rs6662980 in chromosome 1p35).

## 2. Materials & Methods

### 2.1. Samples

Two hundred subjects diagnosed with OCD by DSM IV – TR (APA – DSM, 2000) were identified at a specialty OCD clinic in south India at the National Institute of Mental Health and Neuro Sciences (NIMHANS), Bengaluru and invited to participate after due informed consent. The study was approved by the NIMHANS ethics committee.

Patients were assessed on a detailed clinical proforma employed in the OCD clinic which includes socio-demographic data, age of onset of OCD, duration of illness, duration of untreated illness, detailed history of present illness, presence of common comorbid disorders — including mood disorders, anxiety disorders, tic disorders and other OC spectrum disorders, family history of OCD and major psychiatric disorders, and treatment details. Patients were, in addition, assessed on Yale Brown Obsessive Compulsive Scale (YBOCS) (Goodman et al., 1989b, 1989a), Mini International Neuropsychiatric Interview plus (MINI plus) (Sheehan et al., 1998), and the Clinical Global Impression scale (CGI) (Guy, 1976). Insight was measured using YBOCS item 11 (Goodman et al., 1989a). Diagnosis and associated features were independently confirmed by two clinicians, one of them being a consultant psychiatrist with experience in evaluating persons with OCD. Patients with comorbid psychosis, bipolar disorder and mental retardation were excluded from the study.

Patients underwent routine clinical treatment and follow-up at the OCD clinic. All received pharmacological treatment, which was based on existing standard guidelines (Janardhan Reddy et al., 2017; Koran et al., 2007). Serotonin reuptake inhibitor (SRI) response was defined as reduction of YBOCS > 35% and CGI-I score of 1 or 2 after 12 weeks of treatment (Mataix-Cols et al., 2016). SRI non-responder was defined as reduction of YBOCS < 25% even after 12 weeks of treatment at adequate dose of SRI and CGI-I of 3 and above with, at least two SRIs (Math and Janardhan Reddy, 2007; Pallanti et al., 2002; Shetti et al., 2005). Patients who were partial responders, and who received inadequate trails were excluded from the pharmaco-genetic analysis.

### 2.2 Genotyping

Eight milliliters blood was drawn from the study subjects under aseptic precautions. DNA was extracted using standard techniques (Miller et al., 1988). Genotyping analysis at rs6662980 was done using a commercially available Taqman genotyping assay (Applied Biosystems, assay ID Number 186966013_1). The polymerase chain reaction (PCR) for the Taqman SNP genotyping was carried out as per the manufacturer’s specifications.

### 2.3. Statistical analysis

Fourteen symptom categories of the Y-BOCS checklist (excluding miscellaneous symptoms) were used to generate factors (symptom dimensions). Principal component analysis was performed with ‘varimax’ rotation and an eigenvalue greater than 1 was used to select the number of factors. Factor loadings of greater than 0.50 were considered robust. Univariate analysis of clinical variables (YBOCS total score, insight score, treatment response and the scores obtained on five dimensions) were assessed for genotype association using t-test and chi-square (Fisher’s exact) tests. The Benjamini-Hochberg procedure for false-discovery rate (FDR) correction was used to correct for multiple comparisons. To look for predictors of treatment response, logistic regression was used. The predictor variables included key clinical variables (including age, sex, YBOCS and the factor scores as covariates) and the presence or absence of A/G allele. All analyses were performed using R version 3.6.1.

## 3. Results

### Sample Description

Table 1 shows the clinical and socio-demographic characteristics of the 200 subjects who were included in this study. Of these, 79 were found to have inadequate (<25% reduction in YBOCS or CGI-I>3) response to at least 2 SRI trials on follow-up (median duration = 60 months). Another 98 subjects were found to have adequate response (>35% YBOCS reduction or CGI-I<3) to an SRI treatment. Two subjects were excluded due to lack of follow-up (dropped out of follow-up after initial evaluation), 20 showed partial response (25-35% reduction in YBOCS and CGI-I=3) after one or more SRI trials. Thus for the purpose of comparing the influence of the *SAPAP3* genotype on the response to SRIs, only 177 patients who met the criterion for either response or resistance, were included.

**Table 1.** Clinical Characteristics of the Sample (N=200)

| Characteristic |  | Mean (SD) or n (%) |
| --- | --- | --- |
| Sex | Male | 108 (54%) |
|  | Female | 92 (46%) |
| Age (years) |  | 28.12 (10.41) |
| Age at Onset (years) |  | 19.73 (7.86) |
| Juvenile Onset (<18) |  | 100 (50%) |
| Duration of Illness (years) |  | 8.38 (7.07) (Median=6.5) |
| YBOCS Total Score (Baseline) |  | 25.79 (6.97) |
| CGI-S (Baseline) |  | 4.51 (0.78) |
| Insight |  | 1.29 (0.87) |
| Poor Insight |  | 19 (9.5%) |
| YBOCS Checklist Obsessions (Lifetime) | Contamination | 126 (63%) |
|  | Somatic | 12 (6%) |
|  | Aggressive | 65 (32.5%) |
|  | Sexual | 49 (24.5%) |
|  | Religious | 52 (26%) |
|  | Hoarding | 30 (15%) |
|  | Pathological Doubts | 104 (52%) |
|  | Symmetry | 64 (32%) |
|  | Miscellaneous | 31 (15.5%) |
| YBOCS Checklist Compulsions (Lifetime) | Washing | 127 (63.5%) |
|  | Checking | 104 (52%) |
|  | Repeating | 76 (38%) |
|  | Counting | 22 (11%) |
|  | Collecting | 29 (14.5%) |
|  | Ordering/Arranging | 41 (20.5%) |
|  | Miscellaneous | 46 (23%) |
|  | Mental Compulsions | 44 (22%) |
| **Comorbidities** |  |  |
| Major Depressive Disorder |  | 72 (36%) |
| Dysthymia |  | 8 (4%) |
| Suicidality | no risk | 176 (88%) |
|  | low risk | 17 (8.5%) |
|  | moderate risk | 5 (2.5%) |
|  | high risk | 2 (1%) |
| Generalized Anxiety Disorder |  | 30 (0.15) |
| Panic Disorder |  | 10 (0.05) |
| Social Anxiety Disorder |  | 8 (0.04) |
| Any Anxiety Disorder |  | 43 (21.5%) |
| Tic Disorder/Tourette's Syndrome |  | 10 (5%) |
| Body Dysmorphic Disorder |  | 1 (0.5%) |
| Trichotillomania/Skin Picking |  | 7 (3.5%) |
| Family History of Any Psychiatric Illness |  | 105 (52.5%) |
| Family History of OCD (in FDR) |  | 75 (37.5%) |
| Number of Failed Adequate SRI Trials |  | 1.22 (1.49) (Median= 1;<br>Range 0-6) |
| SRI Resistant<br>(<25% reduction to at least 2 adequate SRI Trials) (n=177) |  | 79 (44.6%) |
| Duration of follow-up at the OCD Clinic (in months) |  | 65.88 (34.65) Median = 60 |

YBOCS – Yale Brown Obsessive-Compulsive Scale; CGI-S – Clinical Global Impression - Severity; SRI – Serotonin Reuptake Inhibitor; FDR – First-Degree Relative OCD – Obsessive-Compulsive Disorder

### Results of the factor analysis

Items in YBOCS checklist which had <10% occurrence in the data were excluded from the factor analysis viz somatic obsessions, counting compulsions, and miscellaneous obsessions and compulsions. Using principal component analysis with “varimax” rotation, 5 factors with eigenvalue > 1.0 were extracted. The loadings in each factor were as follows: symmetry (0.97) and ordering/arranging (0.85) in Factor 1, contamination (0.99) and washing (0.95) in Factor 2, pathological doubts (0.76) and checking (0.98) in Factor 3, aggressive (0.53), sexual (0.63) and religious (0.60) with mental compulsions (0.72) in Factor 4, and hoarding obsessions (0.90) with collecting compulsions (0.97) in Factor 5. The overall model could explain 74% of the total variance.

### Distribution of allelic variants of *SAPAP3*

The Hardy-Weinberg equilibrium was tested and found to have no deviation (Chi-Sq= 1.048, p=0.39). The most common genotype was AA, seen in 148 (74%) of the study population. Both the AG (46/200) and the GG genotypes (6/200) accounted for a total of 26% of the study sample. Hence, we compared those subjects that were AA homozygous (n=148) to those having at least one G allele (n=52), for the association analysis of *SAPAP3* genotype with the OCD phenotype.

### Association with clinical variables

Table 2 shows the results of the univariate analysis. Significantly more contamination obsessions (p=0.008) and washing compulsions (p=0.009) were seen in the G allele carrier group. Using the contamination washing factor score as a continuous measure, a higher score was found in the G allele carrier group (p=0.003). Additionally, a greater proportion of Generalized Anxiety Disorder was seen in those with the AA genotype (or G non carriers) (p=0.007). When the Benjamini-Hochberg procedure was applied with an FDR of 0.10, only contamination/washing was found to survive the correction for significance.

**Table 2.**
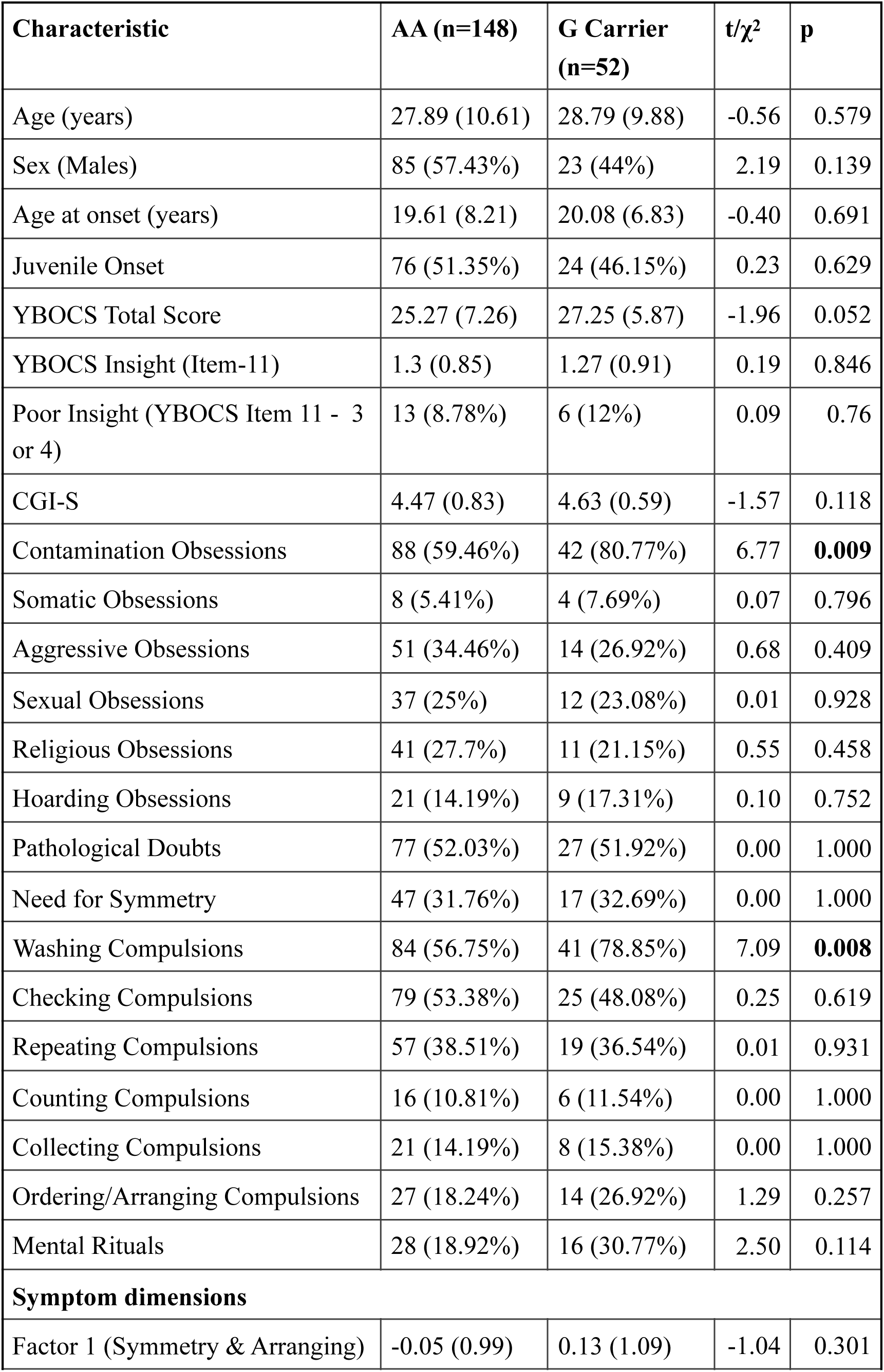
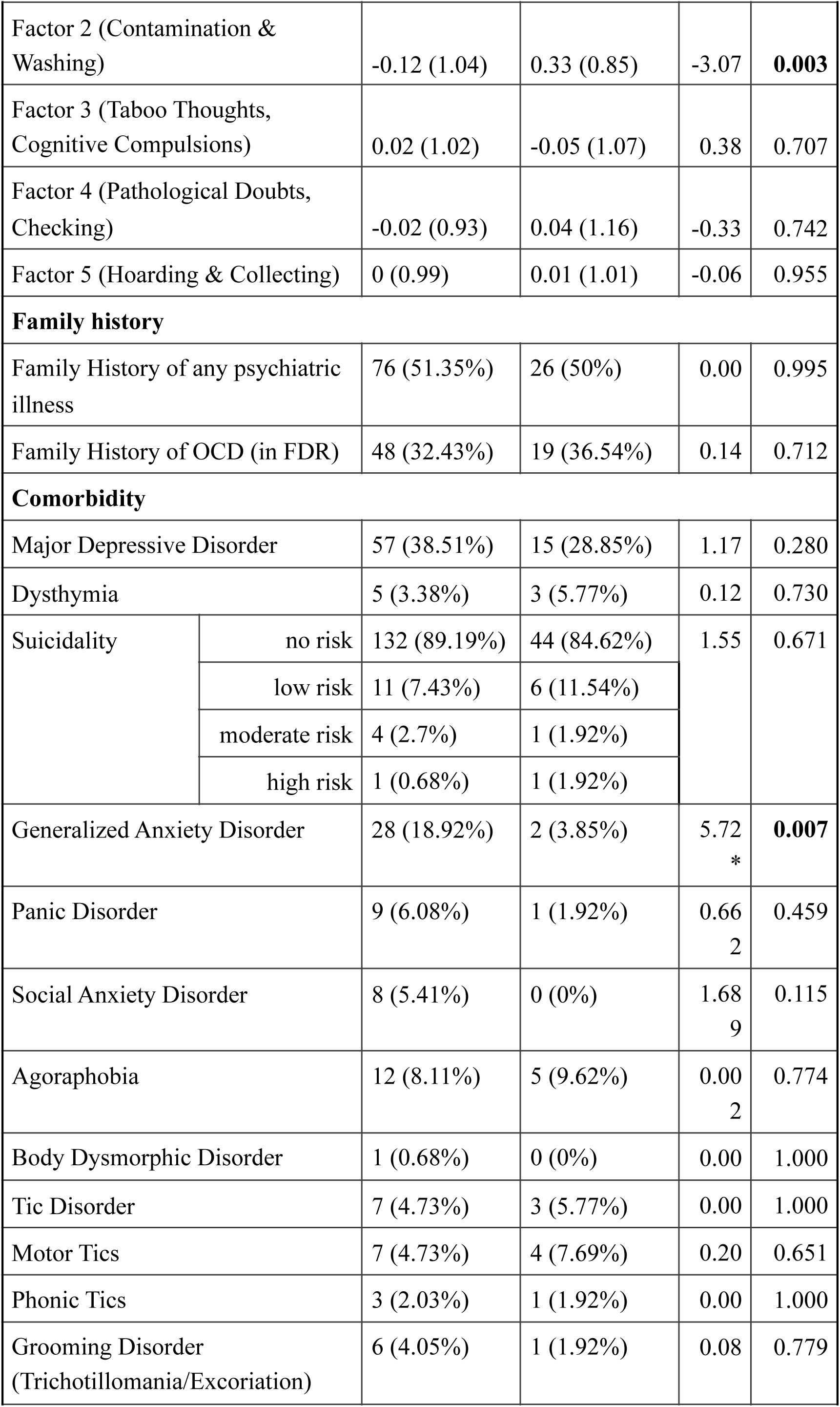
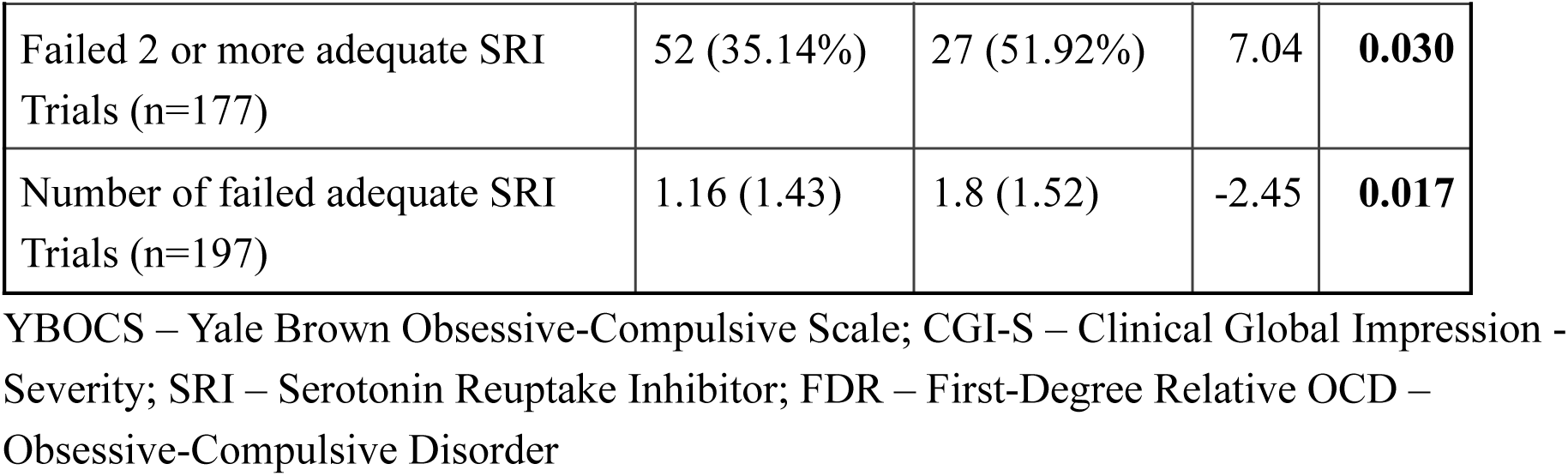
Univariate Analyses of Clinical Characteristics between AA homozygous and G-allele carriers of the *SAPAP3* gene rs6662980 variants (N=200)

| Characteristic | AA (n=148) | G Carrier (n=52) | t/$\chi^2$ | p |
| --- | --- | --- | --- | --- |
| Age (years) | 27.89 (10.61) | 28.79 (9.88) | -0.56 | 0.579 |
| Sex (Males) | 85 (57.43%) | 23 (44%) | 2.19 | 0.139 |
| Age at onset (years) | 19.61 (8.21) | 20.08 (6.83) | -0.40 | 0.691 |
| Juvenile Onset | 76 (51.35%) | 24 (46.15%) | 0.23 | 0.629 |
| YBOCS Total Score | 25.27 (7.26) | 27.25 (5.87) | -1.96 | 0.052 |
| YBOCS Insight (Item-11) | 1.3 (0.85) | 1.27 (0.91) | 0.19 | 0.846 |
| Poor Insight (YBOCS Item 11 - 3 or 4) | 13 (8.78%) | 6 (12%) | 0.09 | 0.76 |
| CGI-S | 4.47 (0.83) | 4.63 (0.59) | -1.57 | 0.118 |
| Contamination Obsessions | 88 (59.46%) | 42 (80.77%) | 6.77 | **0.009** |
| Somatic Obsessions | 8 (5.41%) | 4 (7.69%) | 0.07 | 0.796 |
| Aggressive Obsessions | 51 (34.46%) | 14 (26.92%) | 0.68 | 0.409 |
| Sexual Obsessions | 37 (25%) | 12 (23.08%) | 0.01 | 0.928 |
| Religious Obsessions | 41 (27.7%) | 11 (21.15%) | 0.55 | 0.458 |
| Hoarding Obsessions | 21 (14.19%) | 9 (17.31%) | 0.10 | 0.752 |
| Pathological Doubts | 77 (52.03%) | 27 (51.92%) | 0.00 | 1.000 |
| Need for Symmetry | 47 (31.76%) | 17 (32.69%) | 0.00 | 1.000 |
| Washing Compulsions | 84 (56.75%) | 41 (78.85%) | 7.09 | **0.008** |
| Checking Compulsions | 79 (53.38%) | 25 (48.08%) | 0.25 | 0.619 |
| Repeating Compulsions | 57 (38.51%) | 19 (36.54%) | 0.01 | 0.931 |
| Counting Compulsions | 16 (10.81%) | 6 (11.54%) | 0.00 | 1.000 |
| Collecting Compulsions | 21 (14.19%) | 8 (15.38%) | 0.00 | 1.000 |
| Ordering/Arranging Compulsions | 27 (18.24%) | 14 (26.92%) | 1.29 | 0.257 |
| Mental Rituals | 28 (18.92%) | 16 (30.77%) | 2.50 | 0.114 |
| **Symptom dimensions** |  |  |  |  |
| Factor 1 (Symmetry & Arranging) | -0.05 (0.99) | 0.13 (1.09) | -1.04 | 0.301 |

|  |  |  |  |  |  |
| --- | --- | --- | --- | --- | --- |
| Factor 2 (Contamination & Washing) |  | -0.12 (1.04) | 0.33 (0.85) | -3.07 | **0.003** |
| Factor 3 (Taboo Thoughts, Cognitive Compulsions) |  | 0.02 (1.02) | -0.05 (1.07) | 0.38 | 0.707 |
| Factor 4 (Pathological Doubts, Checking) |  | -0.02 (0.93) | 0.04 (1.16) | -0.33 | 0.742 |
| Factor 5 (Hoarding & Collecting) |  | 0 (0.99) | 0.01 (1.01) | -0.06 | 0.955 |
| **Family history** |  |  |  |  |  |
| Family History of any psychiatric illness |  | 76 (51.35%) | 26 (50%) | 0.00 | 0.995 |
| Family History of OCD (in FDR) |  | 48 (32.43%) | 19 (36.54%) | 0.14 | 0.712 |
| **Comorbidity** |  |  |  |  |  |
| Major Depressive Disorder |  | 57 (38.51%) | 15 (28.85%) | 1.17 | 0.280 |
| Dysthymia |  | 5 (3.38%) | 3 (5.77%) | 0.12 | 0.730 |
| Suicidality | no risk | 132 (89.19%) | 44 (84.62%) | 1.55 | 0.671 |
|  | low risk | 11 (7.43%) | 6 (11.54%) |  |  |
|  | moderate risk | 4 (2.7%) | 1 (1.92%) |  |  |
|  | high risk | 1 (0.68%) | 1 (1.92%) |  |  |
| Generalized Anxiety Disorder |  | 28 (18.92%) | 2 (3.85%) | 5.72<br>* | **0.007** |
| Panic Disorder |  | 9 (6.08%) | 1 (1.92%) | 0.66<br>2 | 0.459 |
| Social Anxiety Disorder |  | 8 (5.41%) | 0 (0%) | 1.68<br>9 | 0.115 |
| Agoraphobia |  | 12 (8.11%) | 5 (9.62%) | 0.00<br>2 | 0.774 |
| Body Dysmorphic Disorder |  | 1 (0.68%) | 0 (0%) | 0.00 | 1.000 |
| Tic Disorder |  | 7 (4.73%) | 3 (5.77%) | 0.00 | 1.000 |
| Motor Tics |  | 7 (4.73%) | 4 (7.69%) | 0.20 | 0.651 |
| Phonic Tics |  | 3 (2.03%) | 1 (1.92%) | 0.00 | 1.000 |
| Grooming Disorder (Trichotillomania/Excoriation) |  | 6 (4.05%) | 1 (1.92%) | 0.08 | 0.779 |
| Failed 2 or more adequate SRI Trials (n=177) | 52 (35.14%) | 27 (51.92%) | 7.04 | **0.030** |
| Number of failed adequate SRI Trials (n=197) | 1.16 (1.43) | 1.8 (1.52) | -2.45 | **0.017** |

YBOCS – Yale Brown Obsessive-Compulsive Scale; CGI-S – Clinical Global Impression - Severity; SRI – Serotonin Reuptake Inhibitor; FDR – First-Degree Relative OCD – Obsessive-Compulsive Disorder

### Association with treatment response

In the univariate analysis shown in Table 2, the G allele carrier group was found to have a greater proportion of SRI-non-responders (p=0.03) and had also received greater number of SRI trials (p=0.017). Table 3 shows the results of multiple regression model, in which we examined predictors of SRI resistance in present group of OCD patients. Prediction accuracy of the overall logistic regression model was 64.7%. We also observed an interaction between presence of G-allele and contamination factor score to predict SRI resistance (B= 1.197, p = 0.00573) (see Figure 1).

**Table 3.** Logistic Regression to Predict Serotonin Reuptake Inhibitor Resistance (N=177)

| Predictor Variable | B | SE | Odds Ratios (95% CI) | p |
| --- | --- | --- | --- | --- |
| (Intercept) | -0.885 | 0.918 | 0.413 (0.066 - 2.466) | 0.335 |
| Age at onset of OCD | 0.002 | 0.033 | 1.002 (0.941 - 1.07) | 0.947 |
| Age at Assessment (years) | -0.022 | 0.025 | 0.978 (0.929 - 1.025) | 0.375 |
| Sex (0=female, 1=male) | -0.056 | 0.361 | 0.946 (0.466 - 1.929) | 0.878 |
| YBOCS Total Score | 0.045 | 0.026 | 1.046 (0.994 - 1.102) | 0.087 |
| Any Anxiety Disorder (0=no, 1=yes) | -0.463 | 0.403 | 0.629 (0.279 - 1.371) | 0.251 |
| Factor 1 (Symmetry & Arranging) | -0.054 | 0.187 | 0.948 (0.653 - 1.367) | 0.774 |
| Factor 2 (Contamination & Washing) | -0.391 | 0.180 | 0.676 (0.472 - 0.958) | **0.030** |
| Factor 3 (Taboo Thoughts, Mental Compulsions) | -0.218 | 0.172 | 0.804 (0.572 - 1.125) | 0.205 |
| Factor 4 (Pathological Doubts, Checking) | -0.108 | 0.173 | 0.898 (0.636 - 1.259) | 0.533 |
| Factor 5 (Hoarding & Collecting) | -0.074 | 0.194 | 0.928 (0.627 - 1.351) | 0.701 |
| Presence of G Allele (0=no, 1=yes) | 0.905 | 0.392 | 2.473 (1.157 - 5.407) | **0.021** |
| **Interaction Analysis** |  |  |  |  |
| Contamination/Washing<br>Factor Score * Presence of G<br>allele | 1.296 | 0.455 | 3.654 (2.761 – 4.547) | **0.004** |

YBOCS – Yale-Brown Obsessive-Compulsive Scale; OCD – Obsessive-Compulsive Disorder

**Figure 1.**
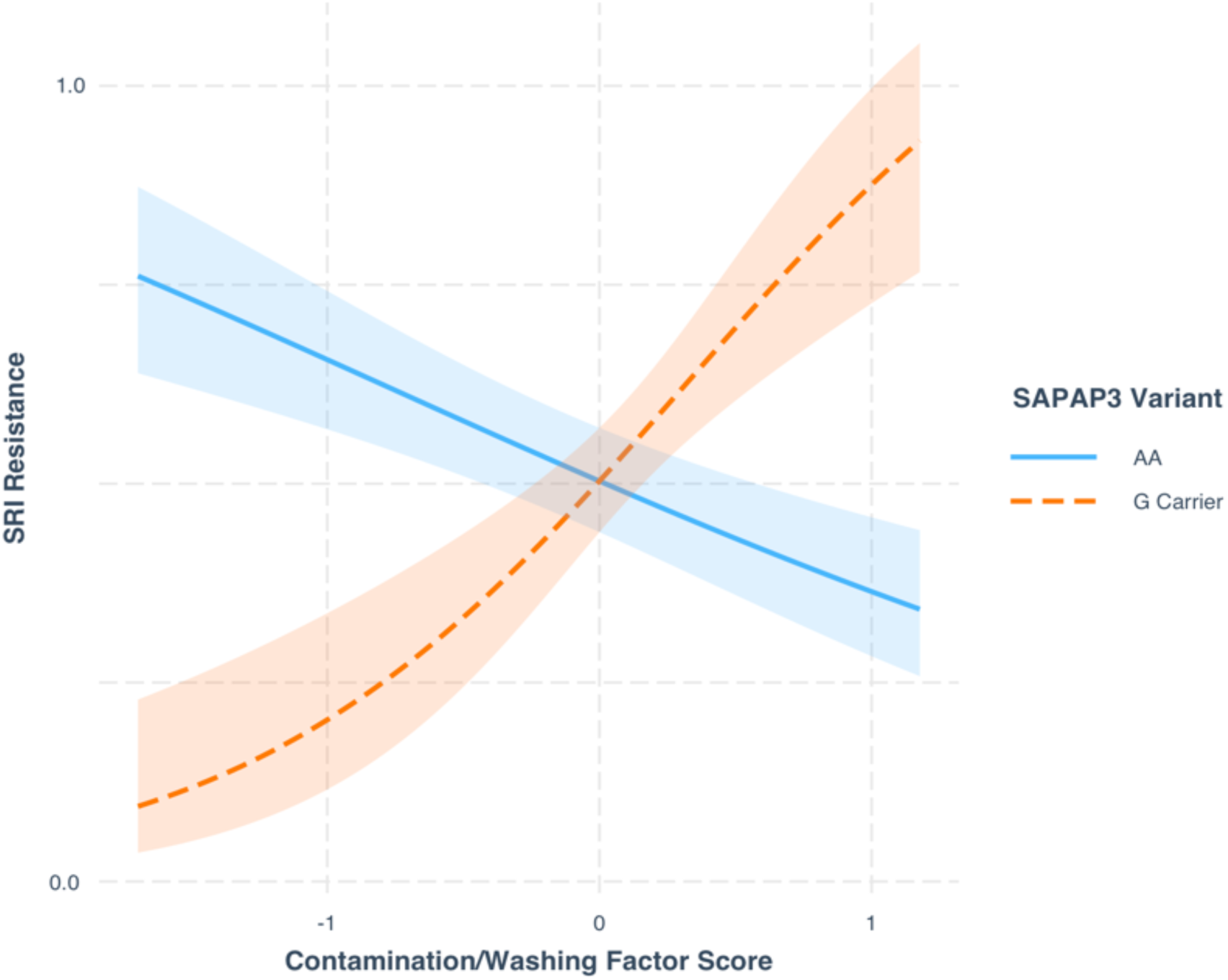
Interaction plot showing differential relationship between contamination factor score and SRI treatment response with and without presence of the G allele in *SAPAP3* rs6662980. *Explanation – In the presence of G allele, those with higher contamination/washing factor score are more likely to be SRI resistant; on the other hand, those with AA allelic variant with higher contamination/washing scores are less likely to have SRI resistance* SRI – Serotonin Reuptake Inhibitor

## Discussion

The current study examined the allelic variants at rs6662980 within *SAPAP3* gene, in a sample of 200 subjects with OCD; and association with clinical characteristics of OCD, including symptom dimensions and treatment response. The two key findings in the study were that the G-allele at this SNP was associated with contamination/washing symptom dimension, and with poor response to treatment with SRIs.

In our study sample, more than one-third of the participants had familial OCD. There was also a greater proportion of those with juvenile onset. Previous studies have found that familial OCD is also associated with more compulsions, greater severity of illness and greater comorbidity (Arumugham et al., 2014; Viswanath et al., 2011). The other characteristics of the sample (symptoms, comorbidity, treatment response) are as in other OCD studies from our center (Taj M J et al., 2018, 2013).

The G allele was found to be associated with the contamination/washing symptom dimension. In light of the evidence from animal models showing excess grooming behaviors after knockout of the gene (Welch et al., 2007), previous studies on *SAPAP3* in humans have specifically examined its relationship with trichotillomania & skin picking disorder (Boardman et al., 2011; Züchner et al., 2009). In our sample, we found only a few subjects with comorbid trichotillomania/skin picking disorder, and hence the association could not be tested. The association between variants in the *SAPAP3* gene and contamination/washing symptoms in our study makes us speculate whether animal grooming behaviors might be analogous to human cleaning/washing behavior, in addition to trichotillomania/skin picking (Ahmari, 2016; Kalueff et al., 2016). Cross-cultural studies on contamination measures have found cleanliness and grooming to co-exist in factor analysis, specifically in non-European samples (Williams and Turkheimer, 2007).

With regards to treatment response, the G allele was found to be associated with SRI treatment resistance. Excessive grooming behaviors observed in *SAPAP3* knockout mice has however been shown to be rescued after treatment with serotonin reuptake inhibitors (Burguière et al., 2013; Welch et al., 2007). Several previous studies have examined the influence of genes on the response to SRIs. A large GWAS which examined treatment response in OCD (Qin et al., 2016), found a locus in *DLGAP1* and multiple other genes in the glutamatergic signaling pathway (Enrichment score = 3.38, FDR=0.00097) to be associated with SRI resistance. Two studies with a candidate gene approach that have examined the association between *SLC1A1* SNPs and treatment response (Abdolhosseinzadeh et al., 2019; Real et al., 2013) have also found an association with treatment resistance. Recently, a study using a polygenic risk score with all the above-mentioned variants (Alemany-Navarro et al., 2019) failed to find any association between the composite score and treatment response in OCD. However, they did not include variants in the *SAPAP3* gene for their estimation of the polygenic risk score.

A key strength of our study includes the use of structured clinical assessments to characterize symptoms, comorbidities and treatment resistance. The use of dimensional-YBOCS (DYBOCS) (Rosario-Campos et al., 2006) ratings could have helped better delineate the phenotype of OCD symptoms, rather than using scores derived from a factor-analysis of the YBOCS checklist. We also assessed treatment response over a naturalistic follow-up, and did not examine response or the lack thereof to specific SRIs. The exclusion of partial responders from our analysis, as we chose to specifically compare “resistant” OCD with responders, could be an additional limitation. Considering that we looked for the association of various phenotypes within OCD (five symptoms dimensions, early-vs adult-onset, comorbidity and treatment response), the sample size of 200 may have been suboptimal. Another limitation is that we did not genotype ancestry informative markers to rule our any effects of population stratification in this study. We have done this previously in another study of OCD from our center and did not find any stratification (Taj et al. 2013).

To conclude, *SAPAP3* rs6662980 genotype was associated with contamination dimension of OCD and SRI treatment resistance in this study. This finding needs to be confirmed in larger genome-wide pharmacogenetic studies (Antypa et al., 2014; Jung et al., 2017) in this population.

## Data Availability

Data associated with findings presented in the manuscript are included in Tables. Any additional data that readers/reviewers may need, may be requested from the corresponding author.

## Acknowledgements

Nil

